# Pervasive transmission of E484K and emergence of VUI-NP13L with evidence of SARS-CoV-2 co-infection events by two different lineages in Rio Grande do Sul, Brazil

**DOI:** 10.1101/2021.01.21.21249764

**Authors:** Ronaldo da Silva Francisco, L. Felipe Benites, Alessandra P Lamarca, Luiz G P de Almeida, Alana Witt Hansen, Juliana Schons Gularte, Meriane Demoliner, Alexandra L Gerber, Ana Paula de C Guimarães, Ana Karolina Eisen Antunes, Fagner Henrique Heldt, Larissa Mallmann, Bruna Hermann, Ana Luiza Ziulkoski, Vyctoria Goes, Karoline Schallenberger, Micheli Fillipi, Francini Pereira, Matheus Nunes Weber, Paula Rodrigues de Almeida, Juliane Deise Fleck, Ana Tereza R Vasconcelos, Fernando Rosado Spilki

**Affiliations:** Laboratório de Bioinformática, Laboratório Nacional de Computação Científica, Petrópolis, Brazil; Laboratório de Microbiologia Molecular,Universidade Feevale, Rio Grande do Sul, Brazil

**Keywords:** COVID-19, E484K, co-infection, P13L, mutation, genomic surveillance

## Abstract

Emergence of novel SARS-CoV-2 lineages are under the spotlight of the media, scientific community and governments. Recent reports of novel variants in the United Kingdom, South Africa and Brazil (B.1.1.28-E484K) have raised intense interest because of a possible higher transmission rate or resistance to the novel vaccines. Nevertheless, the spread of B.1.1.28 (E484K) and other variants in Brazil is still unknown. In this work, we investigated the population structure and genomic complexity of SARS-CoV-2 in Rio Grande do Sul, the southernmost state in Brazil. Most samples sequenced belonged to the B.1.1.28 (E484K) lineage, demonstrating its widespread dispersion. We were the first to identify two independent events of co-infection caused by the occurrence of B.1.1.28 (E484K) with either B.1.1.248 or B.1.91 lineages. Also, clustering analysis revealed the occurrence of a novel cluster of samples circulating in the state (named VUI-NP13L) characterized by 12 lineage-defining mutations. In light of the evidence for E484K dispersion, co-infection and emergence of VUI-NP13L in Rio Grande do Sul, we reaffirm the importance of establishing strict and effective social distancing measures to counter the spread of potentially more hazardous SARS-CoV-2 strains.

**Highlights:**

- The novel variant B.1.1.28 (E484K) previously described in Rio de Janeiro is currently spread across the southernmost state of Brazil;
- The novel variant VUI-NP13L was also identified by causing a local outbreak in Rio Grande do Sul;
- B.1.1.28 (E484K) is able to establish successful coinfection events co-occurring simultaneously with different lineages of SARS-CoV-2.

## 1.1 Introduction

One year has passed since a novel virus, SARS-CoV-2, identified as the etiological agent of a new pneumonia, “Severe acute respiratory syndrome coronavirus 2”, emerged from an outbreak in Wuhan, in the Hubei province in China (Wu et al., 2020; Zhou et al., 2020). It was reported as pandemic by the World Health Organization (WHO) by 11 March 2020, and consequently spread fast to more than 200 countries, causing COVID-19 (*coronavirus disease* 2019) disease. As of January 10, 2021, the WHO has reported 88,383,771 confirmed cases and 1,919,126 deaths globally (https://covid19.who.int/). In Brazil, one of the most affected countries, where no lockdowns or strict movement control of goods and people were applied, confirmed cases surpassed 8 million and deaths mounted to over 200,000 (https://covid19.who.int/region/amro/country/br).

The state of Rio Grande do Sul is the southernmost state of Brazil, with a population of 11.4 million inhabitants. It is bordered by the state of Santa Catarina to the north and northeast, and by Uruguay and Argentina (to the south and southwest and to the west and northwest respectively). The first reported case of SARS-CoV-2 in the state of Rio Grande do Sul was diagnosed on February 29, 2020. While state and city governments issued moderate social distancing policies around mid-March, such as the closing of schools and public gatherings, from mid-April onwards, governments approved relaxing those measures for specific industry or service sectors (Silveira et al., 2020).

On May 11, 2020, state officials implemented the so called “controlled distancing system” (https://distanciamentocontrolado.rs.gov.br/), constituted by a flag system based on the general use of public healthcare, obeying criteria such as occupation of intensive care unit beds and monitoring of expected deaths. The flags and their respective restrictions and flexibilization are to be followed by the economic sector and local municipalities. In this system, the state is divided into 21 geographic regions (named from R01 to R26). By the 11th of August, a more lenient update, called the “shared management system” (https://estado.rs.gov.br/cogestao-regional) was proposed. Local officials could decide whether to follow protocols and rules from the current or a more flexible flag, but only in the case of the “zero-zero rule”, in which there was a steady or decreasing percentage of occupied ICU, availability of hospital beds and expected deaths. The system set maximum levels of use for public spaces, but does not restrict most economic activities including restaurants, gyms, beauty parlors, hairdressers, many non-essential services and other sources of social gatherings. Transportation between cities was also not affected by these restrictions and a high number of active cases was reported throughout 2020. Although the state had already suffered a peak of COVID-19 cases in the winter of 2020, it is currently undergoing a second wave of higher magnitude initiated in late October which has remained in sharp increase until the beginning of January 2021.

After the finding of a new variant of SARS-CoV-2 in Rio de Janeiro (Voloch et al., 2020b), we have made an effort to survey whether this new lineage harboring the E484K mutation could also be found in the second wave of COVID-19 affecting southern Brazil, and we aimed to investigate the presence of other possible lineages circulating in the state. Findings include a new variant which originated in Rio Grande do Sul and simultaneous infection of patients by two different SARS-CoV-2 lineages. We also confirmed the spread of the Rio de Janeiro variant to the south of the country.

## 1.2 Material and methods

### 1.2.1 Sample collection

Samples were obtained from the laboratory diagnostic service of the Molecular Microbiology Laboratory at Universidade Feevale, which receives clinical specimens from suspected patients from 40 municipalities in Rio Grande do Sul. Sampling is performed by local municipal health officers and sent to the Laboratory together with patients’ information including age, gender, clinical signs and possible contact with other suspected or confirmed cases. At the Laboratory, SARS-CoV-2 infection is confirmed using different genome targets from the Charité (E) and CDC (N1/N2) protocols (Corman et al., 2020). In this work, 54% of the analyzed samples were from patients of the Metropolitan Region of Porto Alegre and 46% from the northeast of the state of Rio Grande do Sul. Samples positive for the Charité protocol E gene with a cycle threshold (CT) value between 13 and 20 were chosen for sequencing. A total of 92 samples were selected, 61 of which reported at least one symptom of COVID-19. The age distribution between participants ranged from 10 to 80 years of age (5-year range minimum), and half of the sample pertained to each sex. All samples were collected from November 23 to 30, 2020. This study was approved by the National Committee of Research Ethics and the Institutional Ethical Review Board of the Universidade de Feevale (protocol number: 33202820.7.1001.5348), following Brazilian regulations and international ethical standards.

### 1.2.2 Next-generation sequencing and bioinformatics analysis

We used SuperScript IV First-Strand Synthesis System (Thermo Fisher Scientific, USA) to convert viral RNA to cDNA in our samples. Next, the viral genome was amplified following the Artic Network protocol (https://artic.network/ncov-2019) with the SARS-CoV-2 primer scheme (V3). Sequencing libraries were constructed using the Nextera DNA Flex Library Prep kit (Illumina, USA) and sequenced with the MiSeq Reagent Kit v3 (Illumina, USA). Next-generation sequencing (NGS) data was processed using a bioinformatic pipeline described in our previous study (Voloch et al., 2020a). Briefly, FastQC (v0.11.4) and trimmomatic v0.39 (Bolger et al., 2014) was used for quality control analysis and bad quality reads filtration, respectively. Pre-processed reads were mapped with BWA 0.7.17 software (Li and Durbin, 2009) to the Wuhan-Hu-1 reference genome (NC_045512.2). To further sort, filter reads by mapping quality, and remove duplicates we used samtools v1.10 (Li et al., 2009) and picard v2.17.0 (http://broadinstitute.github.io/picard/). We then performed variant calling using GATK v4.1.7.0 (DePristo et al., 2011) for high-frequency variants and LoFreq v 2.1.5 (Wilm et al., 2012) for low-frequency genetic variants. We annotated the features of each variant with snpEff v4.5 (Cingolani et al., 2012). To obtain the landscape of SARS-CoV-2 population structure, we performed clustering analysis using k-means with high-frequency variants in R. The Elbow method was used to determine the best number of clusters (K). Next, a dataset for each candidate value of K was generated and manually inspected by two independent curators. In addition, by retrieving the set of high and low-frequency variants for each sample, we conducted viral haplotype reconstruction with RegressHaplo (Leviyang et al., 2017).

### 1.2.3 Phylogenomics and SARS-CoV-2 lineage distribution

To determine the relationship of SARS-CoV-2 genomes identified in our study region, contextualized to country level genomes and global data, we built a phylogenomic dataset composed of the 92 genomic sequences described in this work together with 945 complete genomes available from the GISAID (https://www.gisaid.org/) database, described next. First, we selected sequences from the southern Brazilian states of Santa Catarina (n=23), Paraná (n=35), and Rio Grande do Sul (n=136). To evaluate the SARS-CoV-2 lineages between southern genomes and countries sharing borders with Paraná and Rio Grande do Sul states, we included sequences from Argentina (n=55) and Uruguay (n=113). Unfortunately, at the time of data collection (23-12-2020) there were no available genomes from Paraguay, which shares a border with the state of Paraná. We included 180 genomes from the state of Rio de Janeiro containing sequences from C5, which had been observed in our genomes, as well as randomly selected genomes from other Brazilian states (n=201). We also added genome sequences (n=201) from randomly selected countries and collection dates that share the same lineages observed in our 92 samples (B.1.1, B.1.1.143, B.1.1.248, B.1.1.33, B.1.91). In order to improve phylogenetic signal and dating resolution, we discarded sequences obtained from environmental sequences with more than 10% of Ns and with incomplete dates (day missing). The Wuhan-Hu-1 reference genome (Genbank accession number: NC_045512.2) was retrieved from genbank (NCBI) as a representative sequence for the root of the SARS-CoV-2 tree (Rambaut et al., 2020) and completed a final dataset composed of 1037 genomic sequences (**Supplementary Material**). Whole genomic sequences were aligned with MAFFT v7.123b (Katoh and Standley, 2013) using the FFT-NS-2 algorithm. Maximum Likelihood (ML) phylogenetic trees were inferred with IQ-TREE v2.0.3 (Nguyen et al., 2015) using the GTR+F+R2 nucleotide substitution model, which was selected by ModelTest algorithm built in IQ-TREE (Kalyaanamoorthy et al., 2017). Branch support was calculated using 1,000 replicate trees generated by ultrafast bootstrap approximation, also implemented in IQ-TREE (Hoang et al., 2018). Phylogenetic trees and maps were visualized and edited in R version 3.4.2 using packages phangorn (Schliep, 2010), ape (Paradis et al., 2004), ggtree (Yu et al. 2017) and phytools (Revell 2012). Finally, we assessed the clocklikeness of the inferred tree using TempEst v1.5.3 (Rambaut et al., 2016) to generate the root-to-tip regression against sampling dates (correlation coefficient=0.79, R^2^=0.63; detail of newly sequenced samples in **Supplementary Figure S1**).

## 1.3 Results

Whole-genome sequencing of the 92 selected samples achieved, on average, 99,9% of the genome coverage with a mean depth greater than 2000x (**Table S1**). We identified 313 single-nucleotide variants (SNVs) of which 166 were missense, 129 synonymous and 18 in UTRs (**Figure 1A; Table S2**). We also found a total of nine indels targeting orf3a (n = 3), orf6 (n = 2), orf7b (n = 1), orf8 (n = 1), N (n = 1) and 3’ UTR (n = 1) (**Table S3**). Of the eight indels in coding regions, five were frameshift, two corresponded to in frame deletions and one insertion (**Table S3**). Furthermore, we observed an elevated accumulation of variants in the 3’ UTR of the SARS-CoV-2 genome mainly targeting proteins ORF9c, ORF3d, N, ORF3a, and ORF8 (**Table S4**). For the SNVs present in more than 50% of samples, 16 variants were selected corresponding to a mutational signature of the circulating viral lineages in Brazil (**Figure 1A**). According to Pangolin on December 22, sequences from Rio Grande do Sul were classified into five main lineages from 15 localities within the state : B.1.1.248 (86%), B.1.1.33 (6%), B.1.91 (4%), B.1.1 (3%), B.1.1.143 (1%) (**Table S1, Supplementary Figure S2**).

**Figure 1.**
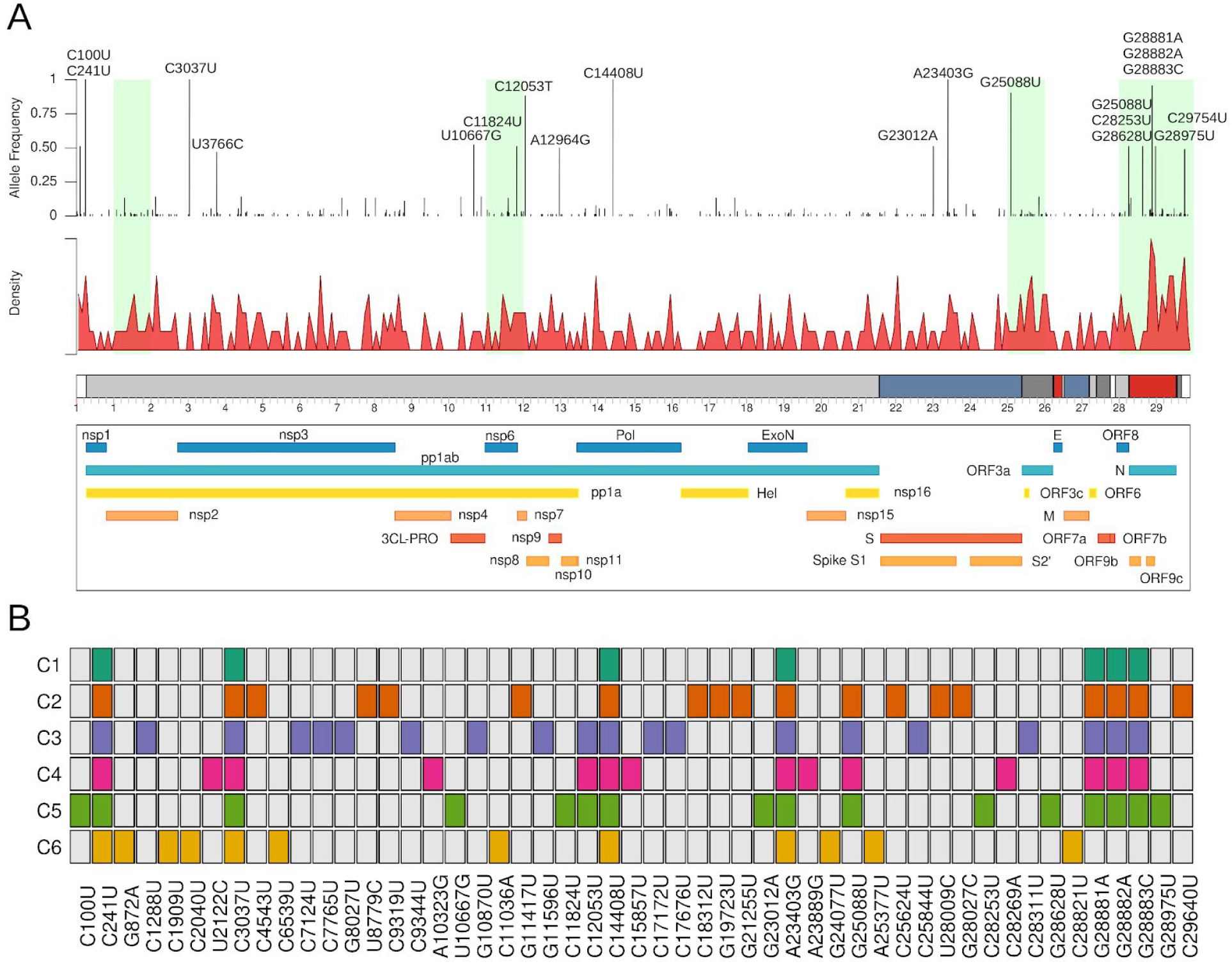
Genomic characterization and variant mapping of SARS-CoV-2 in Rio Grande do Sul, Brazil. A) Distribution of single-nucleotide variants (SNVs) throughout the SARS-CoV-2 genome. Each peak represents the allele frequency of each variant across the 92 samples sequenced. In green, we highlighted the top 5 kilobases with the highest mutation rate. Density plot shows the accumulation of variants in the viral genome. B) Clustering analysis using k-means algorithm with high-frequency SNVs shows six distinct clusters.

We observed a large overlap of minor frequency variants across samples suggesting the possible emergence of novel virus variants. Population structure analysis using k-means algorithm unveiled the existence of six potential clusters (C1 to C6) in our data (**Figure 1B; Table S1**). C1 was composed of 19 samples containing five SNVs also found in other clusters. These samples were classified as pertaining to different lineages such as B.1.1.248, 5 B.1.1.33, 1 B.1.1, and 1 B.1.1.143. However, no discriminant mutations were identified which may indicate the ancient state of the samples in this group (**Figure 1B**). The smallest cluster, C2, contained two sequences from B.1.1.248 that shared 11 exclusive SNVs (five synonymous and six non-synonymous) in non-structural proteins encoded by orf1ab (n = 7), orf3a (n = 1), orf8 (n = 2) and orf10 (n = 1), besides seven fixed mutations considered as a signature of the lineages (**Figure 1B**). The synonymous variant U8514C (F2838F) detected in this group was not previously reported in GISAID, and is first described in this study.

The highest number of exclusive SNVs was shared among samples from the C3 cluster, with 12 mutations occurring in orf1ab (n = 10), orf3a (n = 1) and N (n = 1) in samples from B.1.1.248 and 1 B.1.1 lineages (**Figure 1B**). Based on discriminant mutation profiling in C3, the missense substitution C28311U (P13L) in protein N might represent an important variation since it replaces a Proline by a Leucine at codon 13 in a structural protein. Despite sharing the same set of mutations, these sequences came from 12 unrelated patients from seven different cities in the state. We carried out additional experiments to check whether these viruses were detectable by current standard protocols (Charité and CDC) using all primers and probes. No alterations in amplification from target primers were observed in sequences with the P13L mutation. Thus, all samples were detected regardless of the gene target. Given the emergence of this cluster as a potential novel lineage, we assigned the name VUI-NP13L until Pangolin provides a lineage classification.

In addition, eight samples were assigned to cluster C4 mainly distinguished by indel Δ27900-27903 (F3fs) in orf8 and five other substitutions in orf1ab (n = 4) and S (n = 1) (**Figure 1B**). Samples from C4 harbored a novel missense A23889G (K776R) in Spike, which was not observed in GISAID. Furthermore, sequences clustered in C5 corresponded to the variant under investigation (VUI) derived from B.1.1.248, previously described in Rio de Janeiro (Voloch et al., 2020b). Besides the five SNVs (C100U, G23012A, C28253U, G28628U, and G28975U) reported, we were able to detect two novel mutations in orf1ab, namely U10667G (L3468V) and C11824U (I3853I) both in orf1ab (**Figure 1B**). Different from the first report, the missense variant G23012A (E484K) in S protein, associated with escape from neutralizing antibodies against SARS-CoV-2, occurred in all samples from this group suggesting mutation fixation. Interestingly, the largest number of samples (n = 47) were observed in C5. Altogether, our findings support the hypothesis that this lineage was more recently introduced in Rio Grande do Sul, having probably emerged in Rio de Janeiro. Finally, C6 showed seven distinct substitutions spread across orf1ab (n = 5), S (n = 1), and N (n = 1) (**Figure 1B**). The four samples from this cluster belonged to lineage B.1.91, widely spread in Portugal. B.1.91 was also observed in samples from Porto Alegre in May and July according to GISAID. Moreover, lineages are distributed in clusters as follows: C1 (B.1.1.33=5, B.1.1.248=12, B.1.1.143=1 and B.1.1=1), C2 (B.1.1.248=2), C3 (B.1.1.248=11, B.1.1=1), C4 (B.1.1.248=8), C5 (B.1.1.248=46, B.1.1=1) and C6 (B.1.91=4).

The proposed clusters can also be identified after reconstructing the evolutionary history of the 92 SARS-Cov2 samples. With the inclusion of 957 external sequences, we characterized each cluster clade as the smallest monophyletic group containing all sequences from said cluster (**Figure 2**). As expected, due to its plesiomorphic characteristic, C1 was found to be distributed across the phylogeny and did not form a monophyletic clade. The composition of the C5 clade is of particular interest because it also contains the aforementioned similar sequences from Rio de Janeiro. This corroborates with the hypothesis of a single recent transmission event of the C5 lineage from its origin in Rio de Janeiro to Rio Grande do Sul. With the exception of this isolated event, there is no evidence of association between any other evolutionary lineage and a single locality. This could be attributed to elevated rates of spread due to freedom of host movement despite social distancing measures established (Di Giallonardo et al., 2020). Accordingly, lineage representation in Rio Grande do Sul is similar to that found in Brazil.

**Figure 2.**
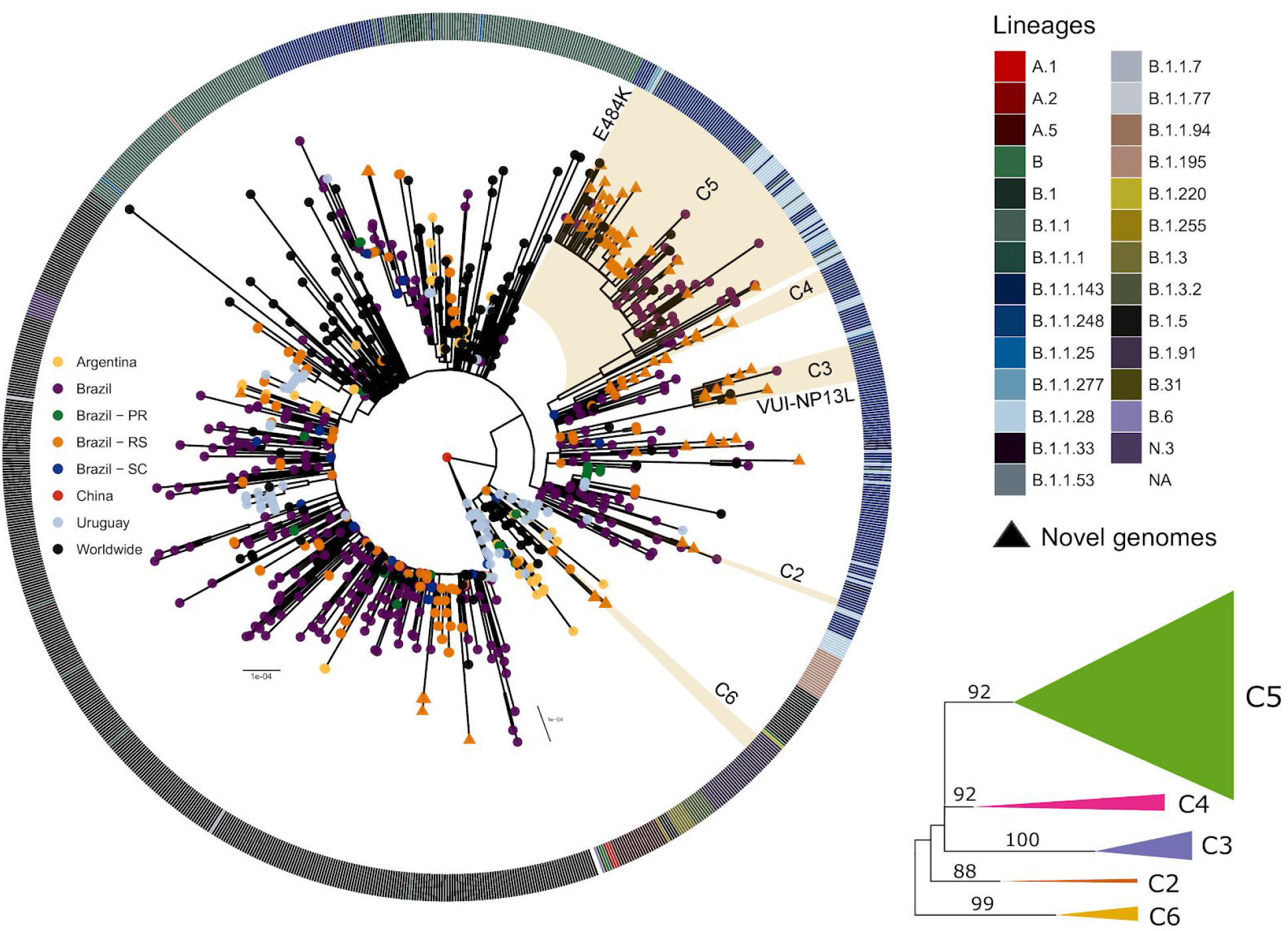
Evolutionary relationships between newly described Rio Grande do Sul samples. A) Maximum likelihood tree inferred with 1037 SARS-Cov2 genomes. Clade highlights indicate the monophyletic grouping for each cluster while the color coded outer ring indicates annotated lineages for each genome based on the Pangolin web application tool. Terminal tip color indicates genome locality by country and southern states of Brazil. In detail, ultrafast bootstrap support for each clade highlighted in A.

We found evidence of SARS-CoV-2 co-infection by distinct lineages in two samples from clusters C5 and C6 (**Figure 3A**). Next, a within-host haplotype reconstruction using low- and high-frequency SNVs was performed to validate the co-infection. Sample 38398 from C5 showed two haplotypes with a frequency of 80% and 20%, respectively (**Figure 3A**). This sample was collected from a patient in their 30’s (5-year range minimum), presenting cough as the prominent clinical sign, that has reported close contact to another confirmed case of COVID-19. The patient fully recovered with no need of hospitalization. Besides the 23 variants called by GATK, we also identified seven low-frequency SNVs using Lofreq with frequencies ranging from approximately 10 to 30% (**Figure 3B**). Twenty sites were discordant between the dominant and the minority haplotype. Part of this discordance corresponded to SNVs that characterized lineages B.1.1.248, B.1.1.33 and those associated with the VUI reported in Rio de Janeiro. Remarkably, the absence of G25088U in the minority haplotype and the presence of the two mutations U27299C and U29148C exclusive of B.1.1.33 supported the classification of B.1.1.248 (Rio de Janeiro) as the dominant haplotype and B.1.1.33 as the minority by Pangolin. Similar results were found in sample 38158 from C6 (**Figure 3A**). The clinical specimen related to this infection came from another patient also in their 30’s (5-year range minimum), presenting headache, cough and sore throat as the main clinical symptoms. The case evolved to cure with no further complications. We were able to identify at least two haplotypes with 83% and 17% of frequency (**Figure 3A**). These haplotypes were classified as B.1.91 and B.1.1.248, respectively. Of note, the dominant haplotype did not harbor 18 alternative alleles of the 29 discordant sites found between both haplotypes in this sample (**Figure 3C**). In both samples, the lineage from Rio de Janeiro (cluster C5) participated as the major and minor haplotype during the co-infection process, respectively. Thus, SARS-CoV-2 co-infection was characterized by the presence of the E484K mutation in S protein supporting the hypothesis of co-infection by two distinct lineages of SARS-CoV-2 in both samples.

**Figure 3.**
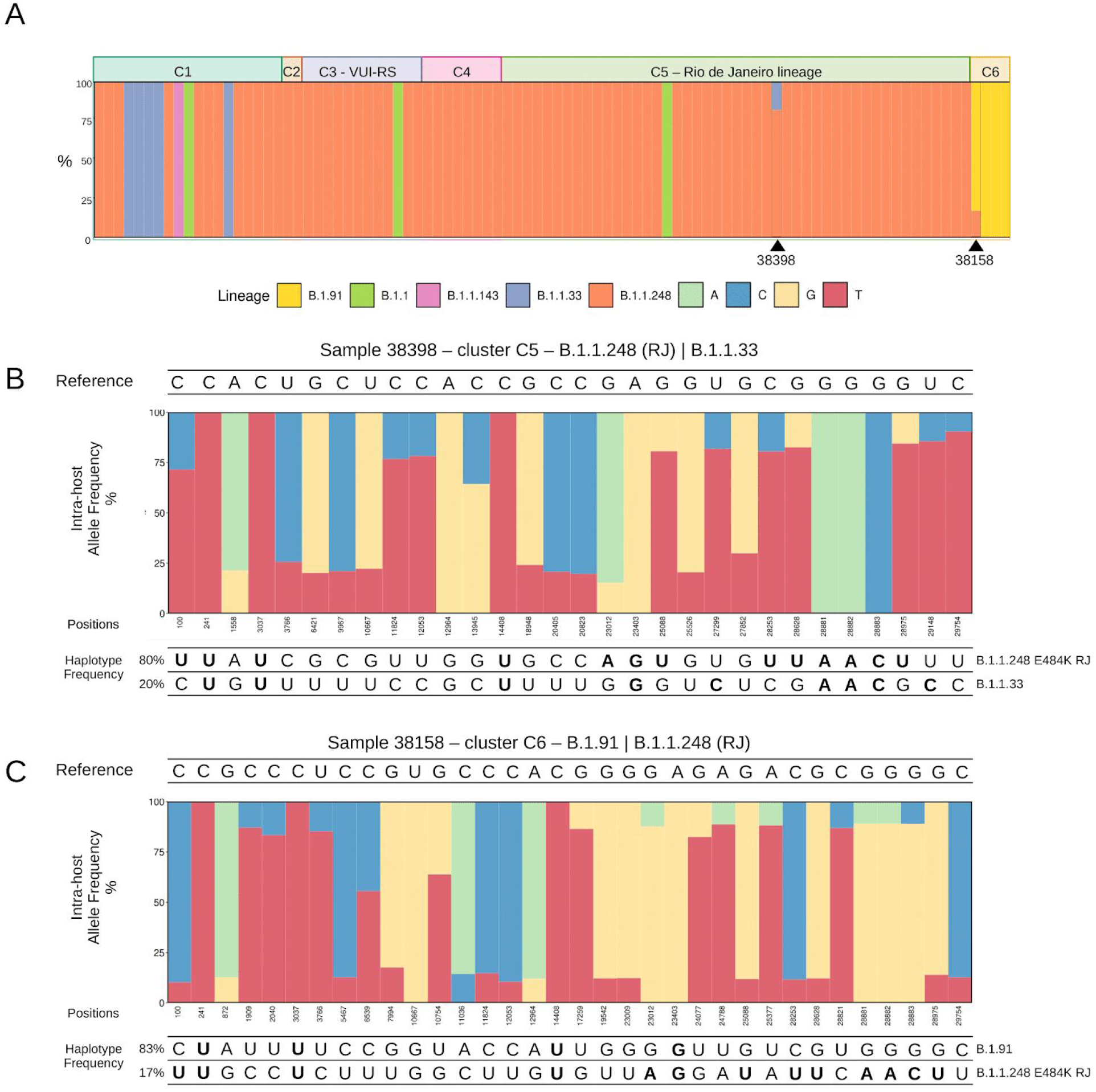
Population structure and intra-host variation. A) Genetic profiling considering clusterization patterns and lineage classification. Bars represent the frequency of SARS-CoV-2 lineages according to Pangolin for each sample using high- and low-frequency variants. The allele frequency pattern observed in samples 38398 and 38158 unveiled two different lineages in both samples. B-C) Intra-host genetic variation analysis in sample 38398 and 38158 showed distinct frequencies of SNVs suggesting the existence of at least two haplotypes. Nucleotides highlighted in bold corresponded to alternative alleles in sites that characterized each lineage.

## 1.4 Discussion

The sharp increase in COVID-19 cases during the so-called “second wave” in Brazil challenges the pursuit of countermeasures and current strategies to keep the epidemic under control. Border regions, such as the state of Rio Grande do Sul, are of particular concern due to the possibility of exportation of viral seeding events to neighboring countries in South America. Indeed, a recent study showed that most of the outbreaks observed in border countries were caused by Brazilian lineages (Mir et al., 2021). As of December 2020, Uruguay and Argentina adopted strict mobility measures by closing the borders for non-citizens to control viral spread. Across the border in Brazil, the metropolitan region of Rio Grande do Sul concentrates the highest number of cases, warning of the possibility of new outbreaks abroad since emergent lineages are constantly being spread across the country. For this reason, in this study we analyzed 92 complete SARS-CoV-2 genomes from the metropolitan region of Rio Grande do Sul to better understand the dynamics, population structure and local transmission chains of the virus in the state. Here, we were able to show that after the introduction of borders restriction by Uruguay and Argentina in late 2020, the viruses found on each side of the borders now separate into different genetic clusters.

Our findings revealed five lineages circulating in the state mainly grouped in six clusters, characterized by co-occurrence of distinct lineage-defining mutations. The absence of exclusive mutations in C1 may suggest an ancient state of samples in this cluster that probably originated the outbreak in the state. C1 was composed of a pool of samples belonging to different independent lineages (most of them classified as B.1.1.33 and B.1.1.248) spread across different segments of the evolutionary tree. Although many genomic surveillance studies focus on mutations in structural proteins, the 11 mutational signatures found in C2 targeting non-structural proteins (orf1ab, orf3a, orf8 and orf10) may represent a valuable set of mutations. For example, mutations in orf8 might have implications in presentation of antigens and viral load (Pereira, 2020). The novel VUI-NP13L variant reported in this study also derived from B.1.1.248 (former B.1.1.28) and harbored an important mutation in the nucleocapsid protein. The P13L mutation also emerged in a cluster from India from stochastic events independent from the Brazilian lineage (Banu et al., 2020). No statistically significant difference was found in the CT values of VUI-NP13L when compared to the other clusters. Structural alterations were also observed in C4 with the presence of a novel missense SNV in Spike. Nevertheless, the impact of such mutation is still unknown. Furthermore, the C3 and C5 clade topological “comb-like” shape reflects a likely rapid exponential growth as a consequence of the epidemic spread (Volz et al., 2013), in contrast with C1 sequences that were distributed throughout several groups, reflected by polytomies, which also suggests possible rapid early spread of these viral clusters (Nie et al., 2020). These nested phylogenetic topologies are predicted in the occurrence of founder effect mechanisms, which in RNA viruses are known to promote rapid changes in genotypes and may also account in phenotypic changes (Bonneau et al., 2001).

Half of the genomes sequenced in our study belonged to the novel B.1.1.28 (E484K) lineage that recently emerged in Rio de Janeiro (clustered in C5). This lineage was also detected in the northeast region of Brazil in the states of Bahia and Rio Grande do Norte (Nonaka et al., 2021). Over the course of the epidemic, we have observed a shift in the prevalence of lineages from B.1.1 (during the beginning of epidemic) to B.1.1.28 (E484K) given its rapid increase in frequency around the country (Resende et al., 2020). In Rio Grande do Sul, the lineage seeding possibly occurred due to a single transmission event, as demonstrated by the phylogenetic analysis. Interestingly, the E484K mutation appears to have arisen independently around the world, as demonstrated by sequences deposited in GISAID. However, only in South Africa (501Y.V2) and Brazil has this substitution been characterized as a lineage-defining mutation always co-occurring with other important substitutions (Tegally et al., 2020; Voloch et al., 2020b). Despite sharing the same variation, it is important to mention that 501Y.V2 harbors a different set of mutations than B.1.1.28 (E484K). Whereas 501Y.V2 is distinguished by eight SNVs targeting Spike protein, the Brazilian lineage was initially described as having five mutations in the UTRs, orf8 and N besides the E484K in S. Since the first report, two more mutations in orf1ab (U10667G>L3468V and C11824U>I3853I) emerged by the end of December (Tegally et al., 2020; Voloch et al., 2020b).

Changes at codon 484 in the receptor-binding domain (RBD) of Spike affected the average binding of convalescent sera (>10-fold) reducing neutralization activity from some individuals (Greaney et al., 2020; Kemp et al., 2020; Piccoli et al., 2020). Escaping the adaptive immune response may be crucial to establishing successful infection even after previous exposure to SARS-CoV-2. Indeed, two recent studies confirmed re-infection cases in Brazil where the second infection was caused by the B.1.1.28 (E484K) lineage (Nonaka et al., 2021; Resende et al., 2021). Finally, this is the first report of co-infection events caused by the simultaneous occurrence of B.1.1.28 (E484K) and other lineages. We have observed that this lineage can participate in the co-infection as either the dominant or minor haplotype. The distinguished pattern in the intra-host frequencies of lineage-defining mutations from B.1.1.28 (E484K), B.1.1.248 and B.1.91 in both cases strongly support this findings. This allowed us to entirely reconstruct the genome of both haplotypes in the co-infection event. Both patients had typical mild to moderate flu-like symptoms with favorable outcomes after disease, no required hospitalization and spontaneous recovery. Although there are a few reported cases of reinfection, the possibility of co-infection by E484K adds a new factor to the complex interaction between immune response systems and SARS-CoV-2 Spike mutations. The putative resistance-associated with E484K may play an important role in cases of co-infection as well as opening a new horizon to investigate changes in the severity of COVID-19 over the course of the infection.

## 1.5 Conclusions

Understanding the phylodynamics, population structure and genomic complexity of SARS-CoV-2 is a powerful approach to establish public health measures aiming to suppress the dissemination of the virus. Nevertheless, limited resources and the few number of samples sequenced in Brazil challenges the continuous monitoring of viral evolution and appearance of novel mutations in the country. The present work demonstrates a pervasive spread of B.1.1.28 (E484K), the possibility of occurrences of co-infection events and emergence of a novel SARS-CoV-2 lineage (VUI-NP13L) across the state of Rio Grande do Sul. The impact of the mutation E484K is still not fully understood; however, its strong association with escaping neutralizing antibodies highlights the necessity for development studies to better establish mechanisms of viral infection. Our results not only increase the number of sequences from the state, but also shed light on an important seeding event between distant Brazilian regions. Yet, further investigation with higher sampling is still needed to access the origin and dispersal of the different variants present in Brazil, including VUI-NP13L. For now, it is remarkable that this new lineage could be found in more than one region in a state supposedly undergoing “social distancing”, showing that these flaccid control measures, in which many activities are being carried out at practically normal levels, may be insufficient to avoid the spread of emerging viral lineages within southern Brazil. We are also conducting *in vitro* experiments including viral isolation and further investigation on the neutralization of VUI-NP13L by convalescent sera antibodies from recovered patients infected with previously reported lineages of SARS-CoV-2. Vaccinated individuals will be investigated as well.

## Data Availability

NGS data generated in our study is publicly available in Gisaid (www.gisaid.org), and the access identifiers are listed in Supplementary Material.

## Figures

**Supplementary Figure 1.**
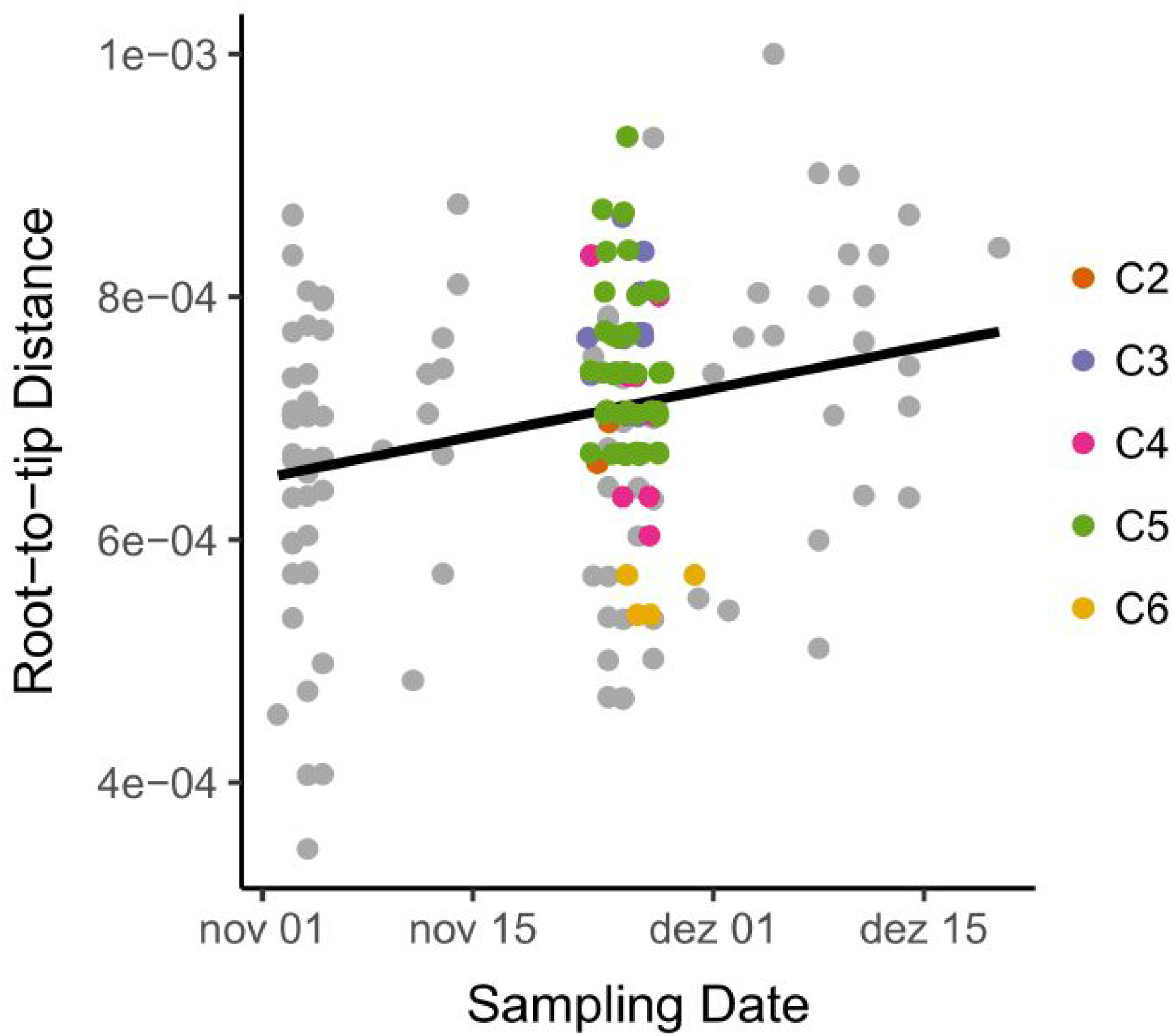
Root to tip regression of SARS-CoV-2 sampled between November and December indicates a clock-like evolution. Colored points represent the newly described 92 sequences from each cluster, while gray points refer to previously published data.

**Supplementary Figure 2.**
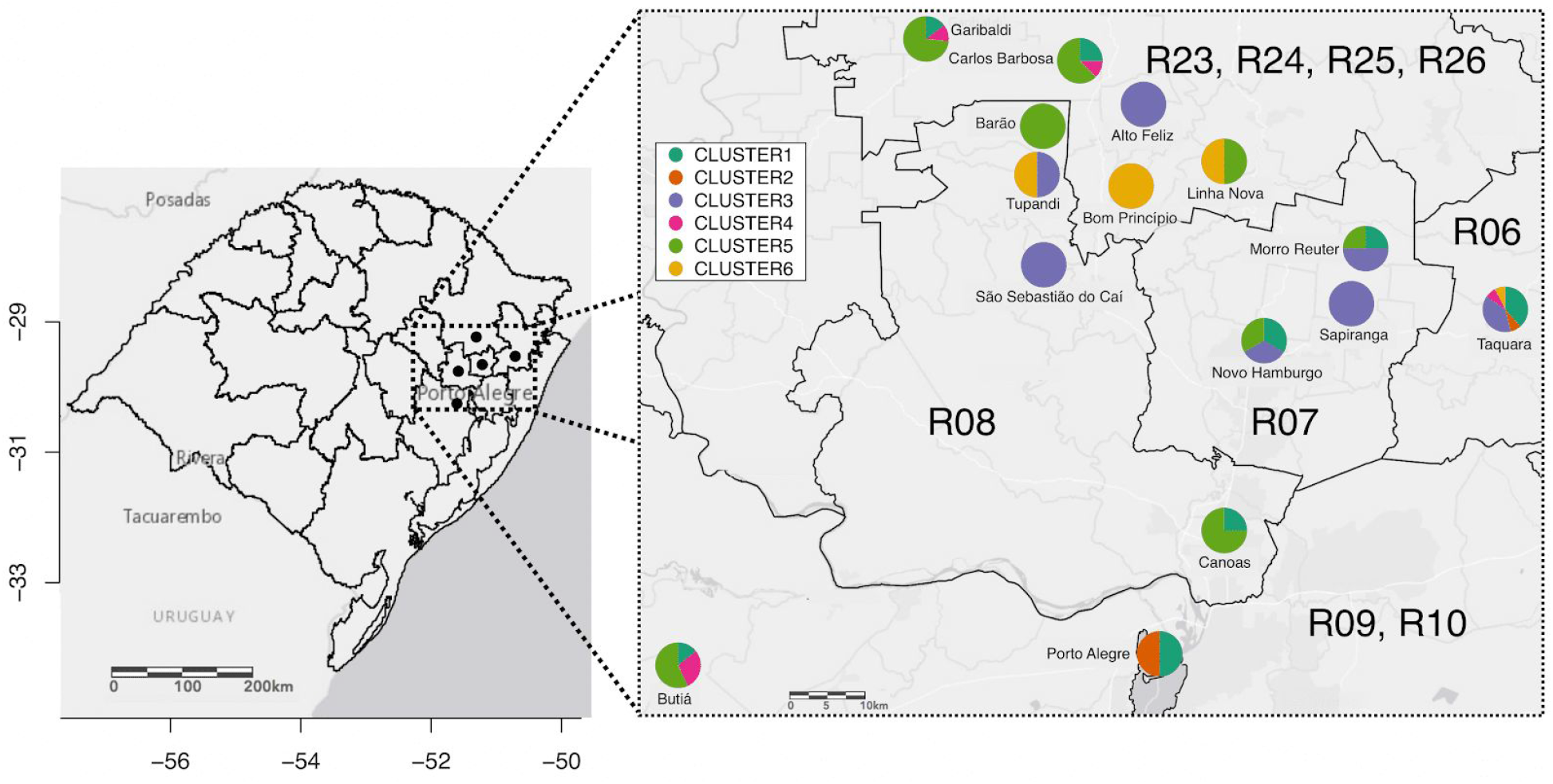
Distribution and frequency of SARS-CoV-2 newly identified Clusters projected on the map of the state of Rio Grande do Sul (30.0346° S, 51.2177° W), with cluster distribution across the regions of the state government’s “controlled ditancing system’’; in A (left) the map of the state of Rio Grande do Sul, and B (right) zoom of 9 regions where the sampling was performed.

## Table

**Table S1 -** Sequencing information and quality control analysis.

**Table S2 -** Single-nucleotide variants identified in the 92 samples from Rio Grande do Sul in Brazil.

**Table S3 -** Insertion and deletion (indel) variants.

**Table S4 -** Per-protein and per-kb mutation rate screening.

## Supplementary Material

Accession ID of the samples sequenced by this study and acknowledgement from the laboratories that deposited sequences in GISAID used in our analysis.

## Acknowledgments

This work was developed in the frameworks of Rede Corona-ômica BR MCTI/FINEP affiliated to RedeVírus/MCTI (FINEP = 01.20.0029.000462/20, CNPq = 404096/2020-4). The study was also supported by Corona-ômica-RJ (FAPERJ = E-26/210.179/2020). A.T.R.V. is supported by Conselho Nacional de Desenvolvimento Científico e Tecnológico - CNPq (303170/2017-4) and FAPERJ (E-26/202.903/20). F.R.S is a CNPq fellow (302668/2018-7). R.S.F.J is a recipient of a graduate fellowship from CNPq. A.P.L and L.F.B are granted a post-doctoral scholarship (DTI-A) from CNPq. J.S.L., M.D., A.K.E.A., K.S. are recipients of CAPES scholarships; F.H.H., B.H., V.G. and M.F. are supported by CNPq grants.

## Author contributions

Data analysis: R.S.F.J, L.F.B, A.P.L, L.G.P.A; Sampling and sample processing: A.W.H, J.S.G, M.D, A.L.G, A.P.C.G, A.K.E.A, F.H.H, L.M, B.H, A.L.Z, V.G, K.S, M.F, F.P, M.N.W, P.R.A, J.D.F; Software R.S.F.J, L.G.P.A; Visualization: R.S.F.J, L.F.B, A.P.L; Funding acquisition: A.T.R.V, F.R.S; Writing: R.S.F.J, A.P.L, L.F.B, A.T.R.V. F.R.S.

